# Using SpliceAI to triage splice-altering variants in 7,220 individuals with rare conditions highlights limitations of the precomputed scores

**DOI:** 10.1101/2025.08.27.25334471

**Authors:** Alexandra C Martin-Geary, Francois Lecoquierre, Susan Walker, Nicola Whiffin, Ruebena Dawes

**Author notes:** Correspondence should be addressed to: Alexandra Martin-Geary or Ruebena Dawes.

## Abstract

**Background:** SpliceAI is a deep learning algorithm that predicts whether genetic variants are likely to affect splicing. Precomputed spliceAI predictions for all theoretical SNVs and small indels were released in 2019, and are used by variant annotation tools such as VEP and dbNSFP to batch annotate large numbers of variants quickly. While the use of precomputed scores vastly reduces the computational cost of retrieving spliceAI predictions, several limitations to their utility have emerged, for which the impact on the prioritisation of likely splice-altering variants in cohort studies has not been established.

**Methods:** We identify annotation and liftover errors present in the precomputed SpliceAI scores by comparing the original gene annotation file with MANE v1.0 Select transcripts. To quantify the impact that these factors may have in variant prioritisation pipelines for diagnostic purposes, we re-analysed variants in 660 genes linked to neurodevelopmental disorders (NDD), identified in 7,221 participants with NDD from the Genomics England 100,000 Genomes Project (100kGP).

**Results:** We find outdated gene annotations and liftover errors impact 8.34% of theoretical SNVs included in the SpliceAI precomputed scores, and affect accuracy of SpliceAI predictions in 34.98% of disease-associated panelApp green genes. In 7,221 Genomics England participants, re-running SpliceAI with optimised parameters results in an increase of 18.2% predicted splice-altering variants compared with variants identified using precomputed scores alone. These variants constitute a diagnostic increase of 11.7% versus the precomputed scores, assessed retrospectively using confirmed diagnoses in 100kGP. In addition we identify a new diagnostic candidate (chr10:129963548:CCGGTGAG:C) in a participant with Ataxia which would be missed by the precomputed scores. To mitigate issues with the precomputed scores we provide ‘SpliceAI-splint’- a command-line tool to allow users to identify variants within a VCF file whose potential splice-altering effects would be missed by the precomputed scores.

**Conclusions:** We find that although the precomputed scores continue to represent a useful resource in the annotation of variants in cohorts with genetic disorders, we find key limitations that can lead to their missing clinically relevant splice-altering variants, and provide a command-line tool to mitigate these issues.

## Background

Genetic variants that affect the recognition of splicing motifs can induce mis-splicing of precursor messenger RNA (pre-mRNA), and are a frequent cause of rare disease(1–3). As such, splicing variants have been a key focus of variant interpretation tool development in recent years. SpliceAI(4) is a well-validated(5–8) deep learning algorithm which predicts whether a genetic variant will alter mRNA splicing. SpliceAI takes up to 5,000 nucleotides (nts) of DNA sequence on either side of a target variant as input, and uses this to predict the likelihood of each nucleotide in the sequence being a donor or acceptor site, in both the reference and when the target variant is introduced. The change in likelihood between the reference and variant sequence (Δ-score) is then calculated for each base. These Δ-scores are further categorised into acceptor gain, acceptor loss, donor gain and donor loss Δ-scores based on whether the site is predicted to be an acceptor or donor site, and whether the variant increases or decreases its strength. By default, the largest Δ-score within 50 nt of the variant is reported as SpliceAI output, within each of the four categories.

The utility of SpliceAI has been greatly enhanced by the provision of precomputed scores for all theoretical SNVs and small indels. These can be used to quickly annotate variants using, for example, the SpliceAI plugin for Ensembl Variant Effect Predictor (VEP)(9). This allows users to rapidly annotate large numbers of variants at a fraction of the computational cost of running SpliceAI directly, and is easily incorporated into pipelines that already use variant effect predictors as part of the annotation process. For many researchers, running SpliceAI naively on large genomic datasets is not feasible without access to large-scale compute resources, and can result in prohibitively high costs when using cloud compute platforms. Consequently, precomputed scores have been widely adopted in both research and diagnostic contexts(10–13).

While undeniably valuable, SpliceAI precomputed scores have several caveats that may limit their accuracy in certain cases. First, SpliceAI relies on a ‘gene annotation’ file containing defined pre-mRNA transcript coordinates which are used when retrieving the DNA sequence surrounding the target variant. For the precomputed scores, this gene annotation file was derived from GENCODE V24lift37, with only the Appris principal transcripts recorded for each gene(4). In the intervening years, largely due to methodological improvements, these annotations have been superseded by the Matched Annotation of NCBI and Ensembl (MANE) project transcript definitions, and in particular their ‘select’ set of transcripts emerging as the variant reporting standard for clinical diagnostics(14).

Second, Δ-scores were only precomputed for ‘small variants’, including all theoretical single nucleotide variants (SNVs), insertions of 1nt, and deletions of up to 4nt in length. This mirrors the fact that larger deletions and insertions are comparatively less common(15,16), and prevents the dataset from being prohibitively large, however users may not be aware that larger indels in their dataset are not receiving SpliceAI predictions when applying the precomputed scores. This is compounded by the fact that variant effect predictors do not differentiate between ‘no predicted splicing impact’, and ‘no data available’ when generating output.

A third limitation of the precomputed scores is that only the maximum Δ-scores within a relatively short distance of 50 nt of the variant are reported. When implementing spliceAI directly, this ‘distance’ parameter can be defined by the user, and in more recent applications, reporting splicing changes a wider distance from the variant (500 nt) has been suggested to improve interpretation of pathogenic splicing variants(7,17–19).

Here, we investigated the limitations of using precomputed spliceAI scores in variant prioritisation pipelines aiming at identifying diagnostic variants in rare disorders. We highlight the reasons for differing predictions between spliceAI precomputed scores, and those obtained directly from the spliceAI command-line tool, and predict the scale of these issues genome-wide. To determine the potential impact of these factors in diagnostic scenarios, we reanalysed SpliceAI predictions for *de novo* variants in NDD associated genes in the Genomics England 100,000 Genomes Project (100kGP)(20).. Building on these findings, we present a framework for the identification of variants warranting reanalysis using updated spliceAI parameters, and provide an open-source tool to enable identification of such variants at scale.

## Materials and Methods

### Identifying LiftOver mapping errors in GRCh38 precomputed scores

To identify regions where LiftOver mis-mapping had occurred in the GRCh38 version of the precomputed scores, we used a custom python script to extract variants from the GRCh38 precomputed SNV vcf for which the REF did not match the sequence in the GRCh38 reference genome. We then used bedtools merge to concatenate overlapping and adjacent positions into a bed file. We applied a padding of 5 nucleotides to each range in the bed file using bedtools slop, and merged these intervals again to produce the bed file to flag regions of reference nucleotide mismatch in the spliceAI precomputed scores. We annotated gene–disease associations within genes with liftover errors using data from Decipher v11.30

### Identifying participants and *de novo* variants

We used genetic data from consenting participants in the Genomics England 100,000 Genomes Project to test our hypothesis. Using the LabKey (v2.9.1) R package inside the Genomics England Research environment we identified an initial cohort of 15,819 consenting probands with a normalised specific disease of “Neurology and neurodevelopmental disorders” from 14,188 families. Of these, 7,220 individuals from 6,522 families (7,073 trios) were present in the *de novo* variant cohort (https://re-docs.genomicsengland.co.uk/de_novo_data/ ) with 428,575 unique *de novo* variants aligned relative to genome build GRCh38, and passing all of Genomics England’s stringency filters.

Variants were filtered to only those in a set of 664 genes associated with NDD, as identified in a meta-analysis of the burden of damaging coding variants in ASD/DD cohorts by Fu *et al*(20). We retained 660/664 genes which had corresponding MANE ‘Select’ transcript annotations. This resulted in a set of 15,244 variants identified in 5,528 trio probands. A VCF file was then generated using these variants, and used for all iterations of the spliceAI and precomputed score analyses.

To identify participants for whom a candidate variant had already been found, we cross referenced our participants, with the Genomics England exit questionnaire and diagnostic discovery tables using RLabKey. In addition to the individuals found in these tables, three participants were identified as newly reported when we returned them to Genomics England, but for which the tables had not yet been updated.

### Running SpliceAI

Precomputed SpliceAI scores were retrieved from the illumina basespace platform (https://basespace.illumina.com/s/5u6ThOblecrh). For all analyses we used the hg38 masked version of the precomputed scores (spliceai_scores.masked.snv.hg38.vcf.gz and spliceai_scores.masked.indel.hg38.vcf.gz).MANE v1.0 .gff annotations were converted to an appropriate format for use in the SpliceAI algorithm using R (see script availability). SpliceAI annotation was performed inside the Genomics England Research Environment using the SpliceAI command-line tool installed in a singularity (v3.8.3) container, the masking option set to “1” and distances set to ‘50’, ‘500’, and ‘4999’.

### Interpretation of genetic variants identified using updated spliceAI parameters

All variants were assessed as primary diagnostic candidates, unless the participant already had an identified diagnostic variant. In these cases variants were assessed on the basis of secondary diagnostic potential.

All genes associated with the identified variants have a known dominant mode of inheritance, as determined by DECIPHER(21). Human phenotype ontology (HPO) terms for each gene were obtained from the human phenotype ontology consortium(22), including frequency of involvement for each term where that information was available. For genes that have not yet been well characterised, possible phenotypic associations were obtained from the literature. These were then compared with the HPO and ICD10 codes recorded for each individual. Any variants which represented a good match for the individual’s phenotype were reported to the participant’s clinical team using Genomics England’s clinical collaboration request system.

To establish the potential for a given predicted splice-altering variant to also have a coding sequence impact we annotated all variants using VEP(9), with the LOFTEE(23) plugin. Any that were within a CDS exon, had a predicted ‘high’ impact, or were annotated as ‘high confidence’ by LOFTEE, and had a consequence of “stop gained”, “frameshift” or “missense” were marked as having a potential coding impact.

### Developing and testing the SpliceAI-splint tool

We built a shell tool (SpliceAI-splint) that takes in variants in VCF format and identifies any warranting reannotation with SpliceAI. To do this, we generated a set of annotation files noting annotation errors in the original SpliceAI gene annotation file versus MANE v1.0 transcripts, alongside the coordinates of LiftOver mapping errors

In addition we ran SpliceAI v1.3 in custom sequence mode (see https://github.com/Illumina/SpliceAI) across the sequences of 19,061 MANE transcripts from GENCODE Version 44 (Ensembl 110), to compute the probability of acceptor and donor splice sites at every nucleotide position along the transcript. Using these reference SpliceAI scores we created an annotation file of locations predicted by SpliceAI to be splice-sites in the reference sequence (donor or acceptor score > 0.02).

SpliceAI-splint uses these annotation files to flag variants in the input VCF overlapping areas with annotation errors, liftover errors, potential long-distance splice predictions, as well as all deletions > 4 nt and insertions > 1 nt.

To confirm that SpliceAI-splint captured all variants identified in our analysis, and establish the proportion of our 15,244 variants that would be annotated for reevaluation with spliceAI, we ran them through SpliceAI-splint within the Genomics England Research Environment.

To establish the expected proportion of variants highlighted that need re-annotation with SpliceAI in different scenarios, we ran SpliceAI-splint on the complete set of 846,357 *de novo* variants aligned to GRCh38 that had passed Genomics England’s stringent filters. We then estimated the expected burden of missed variants per-individual by running SpliceAI-splint on five randomly selected whole-genome small variant (<50bp) VCFs (GRCh38) called using delivery version V4, that had previously been used by Genomics England in their own interpretation pipeline.

We additionally applied spliceAI-splint to two further variant sets outside of The National Genomics Research Library, (1) the GRCh38 ‘genome in a bottle’ v4.2.1 and (2) the 1,000 Genomes Project biallelic SNV and indel VCF, obtained from the ftp site (https://ftp.1000genomes.ebi.ac.uk/vol1/ftp/data_collections/1000_genomes_project/release/20190312_biallelic_SNV_and_INDEL/).

## Results

### SpliceAI precomputed scores contain outdated gene annotations or liftover errors in >35% of genes

We assessed the potential impact of changes in transcript annotations on the accuracy of precomputed SpliceAI scores by comparing the original gene annotation file in the SpliceAI github repository (derived from GENCODE V24lift37 Appris principal transcripts), with a gene annotation file derived from MANE v1.0 transcript definitions.

We interrogated four scenarios in which we reasoned changes to gene annotations could alter reported SpliceAI scores (Figure 1A). First, if a gene is not included in the gene annotation file no predictions will be generated for variants within that gene. Second, SpliceAI includes an option to ’mask’ scores, meaning increases in strength of annotated splice-sites or decreases in strength of unannotated splice-sites will be set to zero. This has the benefit of limiting reported splicing predictions to those likely to be clinically relevant, however if annotated splice-junctions (such as alternative exons) differ between gene annotation files, masking can alter reported predictions.

**Figure 1:**
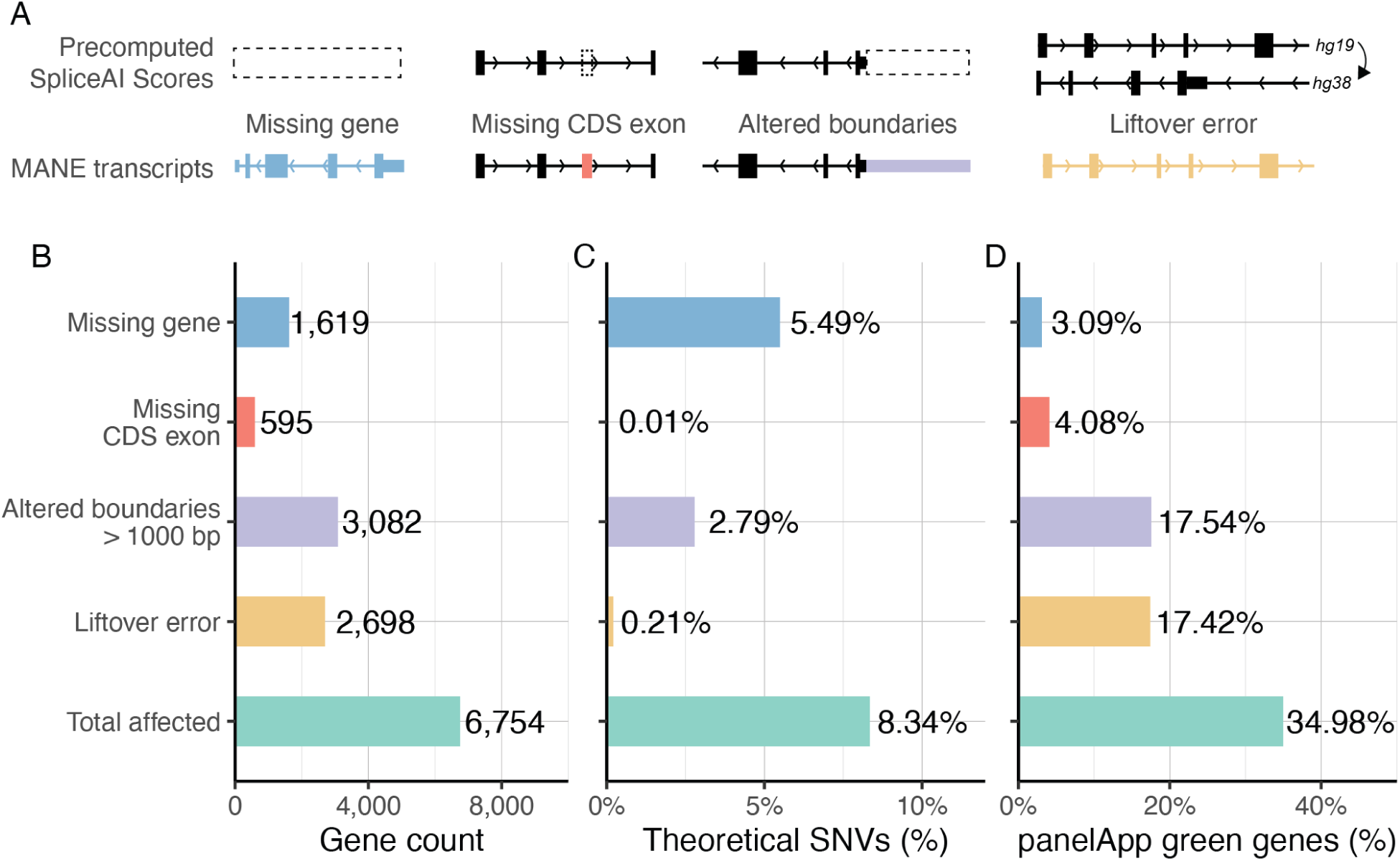
Outdated annotations and liftover errors impact SpliceAI precomputed scores in clinically relevant genes. **A.** Diagram showing instances where annotation issues in the SpliceAI precomputed SNV file may lead to altered predictions for genetic variants. Black dotted lines indicate changes in gene annotation files. **B**. The total number of MANE v1.0 transcripts that differ from the GENCODE V24lift37 transcripts used for the SpliceAI precomputed score calculations. **D**. The percent of all theoretical SNVs in the SpliceAI precomputed scores that overlap each annotation issue. **C.** The percent of all PanelApp genes which have each or any annotation issue in the SpliceAI precomputed scores.

Third, changes to the transcript boundaries reported in the gene annotation file can alter reported predictions in two ways: (1) If a variant falls outside of the annotated transcript coordinates reported in the gene annotation file, no SpliceAI prediction will be generated, (2) only genetic sequence within the annotated transcript boundary will be considered when generating splicing predictions, so even a variant within the annotated transcript coordinates can have an altered prediction if the transcript boundaries have changed.

Finally, we observed that REF alleles in GRCh38 precomputed scores occasionally did not match the sequence of the reference genome. These errors likely derive from faulty liftover of GRCh37 precomputed scores to GRCh38, and result in absent or incorrect splicing predictions. We found that 4.4 million out of 3.4 billion precomputed SNVs in GRCh38 (0.13%) had an incorrect reference allele indicating that liftover had resulted in a mapping error at that site.

Notably, 101 genes (0.49%) have incorrect REF alleles for > 10% of their nucleotides, with 89 (0.44%) having incorrect REF alleles for more than 50% (Supplementary table 1), including a number of genes involved in disease such as *PEX11B* (peroxisome biogenesis disorder), *RBM8A* (thrombocytopenia-absent radius syndrome), GDF2 (hereditary haemorrhagic telangiectasia) and *RBP3* (retinitis pigmentosa), making the precomputed SpliceAI scores unusable for these genes. The 10 genes for which this issue was most severe had incorrect REF alleles for an average of 0.77% of their entire length (Supplementary table 1). In extended genomic regions where incorrect reference alleles indicate liftover mapping errors with GRCh38, we expect ¼ of bases will have the correct reference nucleotide by chance, aligning well with the proportion of correct REF alleles for these genes. To ensure we could flag any variants in regions of liftover error which nonetheless have the correct REF allele we added 5 nucleotides padding to any position with a reference mismatch. We then merged all these padded intervals to identify regions where LiftOver mapping errors limit the accuracy of precomputed scores.

Overall, 6,754 unique genes in MANE v1.0 (35.4% of all genes) are impacted by at least one of these four annotation errors in the precomputed SpliceAI scores (Figure 1B). Of the 19,061 genes in MANE v1.0, 1,619 (8.5%) were entirely missing from the original gene annotation file, with a further 595 (3.1%) genes missing at least one CDS exon that is annotated in MANE v1.0, meaning the score of any variant impacting these newly annotated splice-sites will be set to 0 in the precomputed masked scores. Furthermore, 3,082 genes (16.2%) had altered boundaries of > 1,000 bp at one or both ends and 2,698 genes (14.2%) overlapped at least one liftover mis-mapped region (Figure 1B). Genomic regions containing missing genes, missing exons, altered transcript boundaries and liftover errors in the original gene annotation file affect 8.34% of the 3.4 billion precomputed SNVs in GRCh38 in total (Figure 1C).

To establish the extent to which these problematic regions overlap with clinically important regions of the genome, we intersected them with genes that have strong disease associations recorded in panelApp(24), Genomics England’s expert curated knowledgebase of gene:disease associations (Figure 1D). Overall, 1,416 (34.98%) ‘green’ panelApp genes overlapped with at least one annotation issue identified in the precomputed scores, and 146 of these genes have >50% of all theoretical SNVs impacted by annotation or liftover errors in the precomputed scores (Supplementary Table 2).

### Updating the gene annotation file supplied to SpliceAI identifies predicted splice-altering variants missed by precomputed scores

To determine the extent to which the issues outlined here may negatively impact the interpretation of variants in a real-world context, we performed an example analysis using participants from the Genomics England 100,000 Genomes Project, focusing on a set of 664 NDD-linked genes(20). In a cohort of 7,221 100kGP participants recruited with NDD related phenotypes, we identified 15,244 *de novo* variants in NDD-linked genes (5,528 probands) including 14,885 small (SNVs, 1nt Insertions, deletions <= 4nt) and 359 larger variants (>1nt Insertions & >4nt deletions). Of the 14,885 small variants, 165 were predicted to be splice-altering using SpliceAI (masked) precomputed scores, with the recommended threshold for high recall (max Δ ≥ 0.2). Reannotation of all small variants using SpliceAI (masked) scores (max Δ ≥ 0.2) with updated MANE v1.0 transcript annotations identified 167 *de novo* variants predicted to be splice-altering. Between the two sets of predictions, 164 were annotated with max Δ ≥ 0.2 in both sets; 1 was exclusively annotated in the precomputed scores; and 3 were only identified using spliceAI directly with updated transcript annotations.

We investigated how an updated gene annotation file had resulted in SpliceAI score changes for these three variants. For one variant (chr15:92974877 A/G, max Δ = 0.99), the gene (*CHD2*) was absent from the original gene annotation file used to generate precomputed scores. The second variant (chr11:61780797 G/C, max Δ = 0.74) impacted an exon in MYRF which was not in the original gene annotation file, which meant the score had been masked in the precomputed set. The final variant was identified in the gene *DNMT3A* (chr2:25276150 C/A, max Δ = 0.21), and was missed as the gene annotation file included a short isoform for this gene, and the variant falls outside of this isoform’s boundaries. This isoform excludes 23.3% of the CDS of the MANE isoform, as well as 8.6% (13/151) of pathogenic/likely pathogenic ClinVar variants reported in *DNMT3A*. The variant that was only identified using precomputed scores had a max Δ > 0.2 in only one of the two overlapping genes that it was annotated with reference to. This gene is a novel protein coding gene that is currently poorly characterised, and as such was not included in the MANE gene set. These variants highlight the importance of considering the gene annotation file supplied to the SpliceAI algorithm, as it determines the genetic sequence used to generate predictions.

### Rerunning SpliceAI with optimised parameters increases the number of predicted splice-altering variants by 18.2% relative to the precomputed scores

In addition to the three variants newly identified due to gene annotation file changes, we identified 12 longer indels (deletions ranging between 5 and 17 nt, and insertions between 2 and 5 nt) with max Δ ≥ 0.2. Of all >1 nt insertions and >4 nt deletions present in our set of 15,244 variants 4.7% (12/254) were identified as likely splice-altering (max Δ ≥ 0.2). This is in contrast with 1.1% of SNVs/small indels identified as likely splice-altering (168/14,990 max Δ ≥ 0.2, Fisher’s 2-sided *P*=4.942×10-5). This highlights the benefit of running spliceAI directly over longer indels that are not included in precomputed scores. Overall, when rerunning SpliceAI directly with an updated gene annotation file, 6.2% (952/15,244) of *de novo* variants in our NDD cohort changed their max Δ-score. We found a 9.1% increase in variants annotated as likely splice-altering (max Δ>= 0.2), relative to the precomputed scores (Figure 2A).

**Figure 2.**
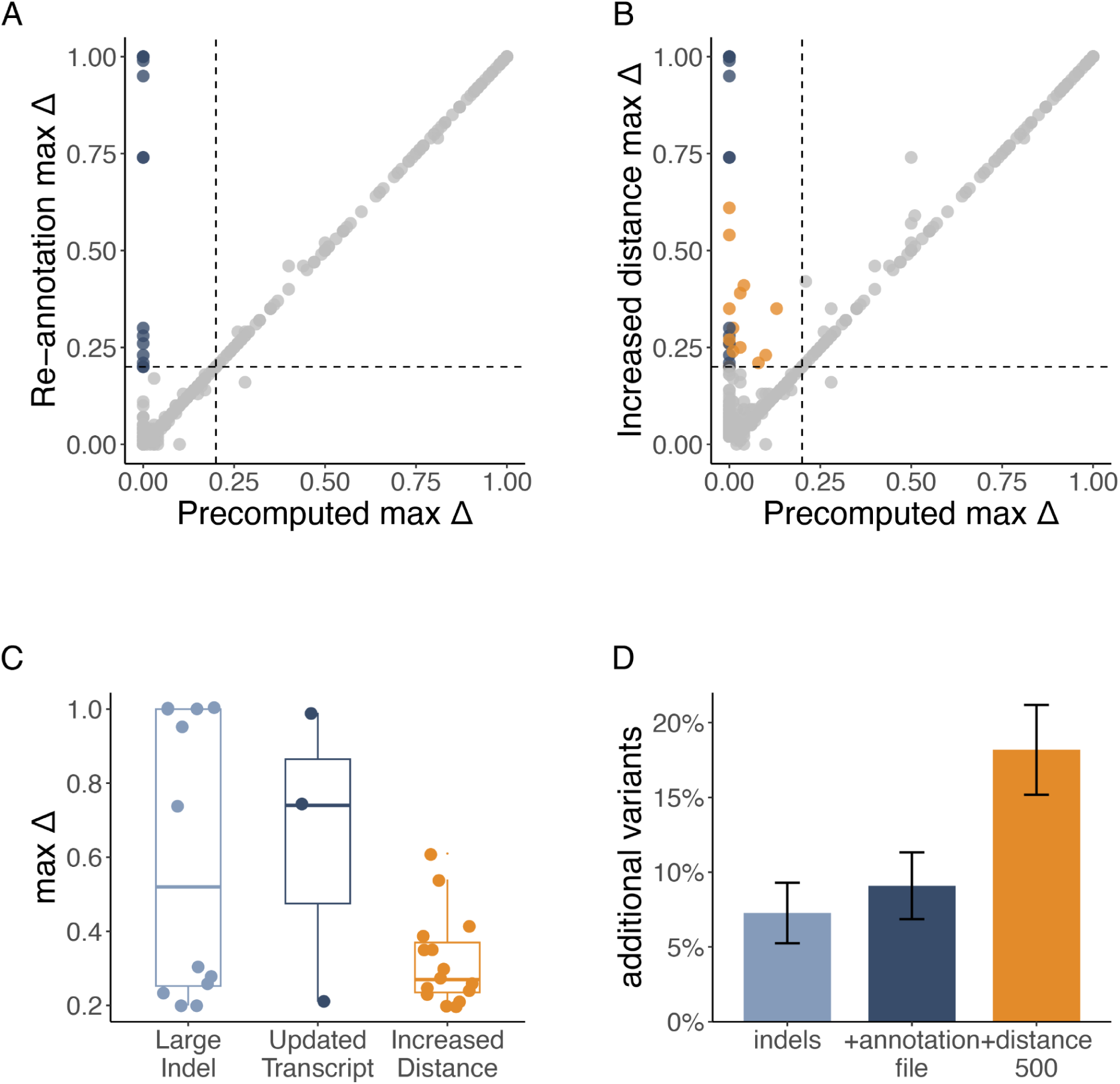
Re-running SpliceAI with optimised parameters identifies additional predicted splice-altering variants. **A.** Updating transcript annotations to MANE v1.0 identifies an additional 15 predicted splice-altering variants (max Δ ≥ 0.2; shown in blue) versus when using the spliceAI precomputed. **B.** Increasing the distance parameter to 500 nt identifies another additional 15 predicted splice-altering variants (max Δ ≥ 0.2; shown in orange). **C.** Re-annotation max-Δ of 30 variants missed using precomputed scores, split into the reason they were missed. **D.** Proportion increases in the number of variants with max Δ ≥ 0.2 due to including indels, updating the annotation file and increasing distance to 500, calculated relative to the number of variants with max Δ ≥ 0.2 identified using precomputed scores (n = 165). Error bars show the standard error (SE) of the proportional increase in variants, calculated relative to the baseline of 165 precomputed variants.

We next increased the distance parameter of SpliceAI to 500 nt, to identify any variants predicted to alter the strength of a splice-site located within 500 nt of the target variant (rather than the default 50 nt). By updating this parameter we are able to detect variants that create cryptic splice-sites at larger distances, or weaken annotated splice-sites by modifying more distal splicing regulatory elements rather than the splicing motif itself.

Using this increased distance we identified a further 15 variants with max Δ ≥ 0.2, including one additional deletion that exceeded the size considered in the precomputed scores (Figure 2B, Supplementary table 3). To determine the potential impact of further increases to distance we repeated this step using the maximum distance of 4,999nt- however no additional variants with max Δ ≥ 0.2 were identified. In fact the most distal predicted change to a splice-site was at 155nt from the target variant, even when allowing for predictions up to 500nt away (Supplementary table 4).

Upon inspection, three of the 15 variants detected by increasing the distance parameter were predicted to increase the strength of an existing annotated splice-site, despite the masking option being activated. As the in-built masking option only operates on the closest splice-junction to the variant, running SpliceAI with an increased distance parameter means predicted changes to more distal splice-sites are not masked correctly. In addition, these variants generally had lower max Δ scores than variants identified with the distance parameter set at the default 50 (Figure 2C). Therefore, while increasing the distance metric increases the number of predicted splice-altering variants, these variants may require higher levels of manual review to ensure that predicted splicing effects are clinically relevant.

In total, rerunning SpliceAI on all variants with an updated gene annotation file and the maximum search distance set to 500 nt resulted in a 18.2% gain in variants (N=30) annotated as likely splice-altering, relative to the precomputed scores (Figure 2D, Supplementary figure 1).

### Identification of existing diagnostic splice-altering variants in NDD participants in 100kGP

We assessed the potential diagnostic increase represented by running SpliceAI with updated gene annotations and optimised parameters, over only the precomputed scores, by assessing ‘known’ candidate diagnostic variants in our NDD participants in 100kGP. We focused on the 14,030 of the 15,244 that were annotated as synonymous, intronic, or UTR exonic, to avoid including candidate diagnoses made due to coding rather than splicing impact. In total, 67 variants with max-Δ>= 0.2 identified by our SpliceAI re-analysis are already recorded in 100kGP as the candidate variant in ‘solved’ families, versus 60 in the precomputed scores, a diagnostic increase of 11.7%.

These additional seven candidate diagnostic variants would have been missed by approaches screening for splice-altering variants using the precomputed scores, as four were larger indels, one was in a gene missing from the precomputed scores (*CHD2*), and two only reached the delta-score threshold when we increased the distance parameter. These variants highlight the importance of running SpliceAI with appropriate gene annotations and distance cutoffs to avoid missing potentially diagnostic splice-altering variants.

### Interpretation of ’rescued’ predicted splice-altering variants identifies novel candidate diagnostic variants excluded from precomputed scores

We additionally assessed variants predicted to be splice-altering in our reannotation as potential novel candidate diagnostic variants. We assessed all of our newly identified variants as potential diagnoses, based on the gene’s phenotypic match with the participants phenotype, and whether the participant already had a candidate diagnostic variant in another gene (Supplementary Table 3, Supplementary Figure 2). We found one variant to be a good candidate. This variant was an intronic 7 nt deletion predicted to decrease the strength of the acceptor of exon 2 of *EBF3* (Δ 0.2), a gene associated with *Hypotonia, ataxia, and delayed development syndrome*(25–27) (chr10:129963548:CCGGTGAG:C). Three of the Top-4 SpliceVault events for this acceptor are out of frame(17), making loss-of-function for this allele probable, however RNA studies would be needed to be conclusive about the splicing phenotype. This variant was identified in an individual recruited with a developmental disorder of speech and language and ataxic gait, making the gene a strong match for this individual’s phenotype.

### Developing a tool to identify variants which require updated SpliceAI predictions

We developed a framework to identify variants which would likely benefit from re-annotation with the SpliceAI command-line tool rather than relying on precomputed scores. We used regions identified to differ between the original SpliceAI gene annotation file and MANE v1.0 transcripts (Figure 1B) to create an annotation file covering regions where changed transcript annotations limit the accuracy of precomputed scores.

Additionally, we noted that all variants we identified using an increased distance parameter were predicted to alter the strength of a splice-site already predicted in the reference genomic sequence, with ref splice-site scores ranging from 0.02 to 0.94 (Supplementary Table 4). This stands to reason, as a variant affecting the strength of a splice site > 50 nt away by definition cannot be creating a core splice-site motif, but must be modifying the strength of one which is already present in the reference sequence. We reasoned we could use this principle to identify variants most likely to benefit from reannotation with an increased distance parameter.

We used the following criteria to identify candidate variants which could benefit from re-annotation:

a. Insertions larger than one nt or deletions larger than four nt
b. Are located within regions identified as LiftOver mapping errors in GRCh38
c. Are located within a gene missing from the original SpliceAI gene annotation file
d. Are located within an exon included in a MANE v1.0 transcript but not the original SpliceAI gene annotation file- including 20 nt padding surrounding the exon
e. Are located within 1000 nt of transcript boundaries, where those boundaries differ by > 1000 nt between MANE v1.0 annotations and the original SpliceAI gene annotation file
f. Are more likely to impact the strength of splice sites > 50 nt away. Informed by variants identified with D500 (Supplementary Table 4), we flag variants within 50-160 nt of a splice-site predicted by SpliceAI to be a splice-site in the reference sequence (donor or acceptor score > 0.02)

Using this schema we developed ‘SpliceAI-splint’ an open-source tool for the re-annotation of variants that are likely to be impacted by one of the criteria set out above. This tool takes in a variant file in VCF format, and flags any variants meeting criteria a-f. Using this tool, all variants identified in our analysis as being missed in the precomputed scores were flagged for reannotation, and 4,473 (29%) of the *de novo* variants, are flagged for reannotation in total. We additionally ran SpliceAI-splint on 5 randomly selected individual VCF files in 100kGP, Genome in a bottle variants, and 1000 Genomes variants for which 12.94-14.62% of variants were flagged for reannotation, reducing the overall burden of re-annotating large datasets by ∼71 - 87% compared to re-running SpliceAI across all variants (Supplementary Table 5).

**Table 1:** The total number, and percent of all variants identified as requiring re-annotation (red categories) using the *SpliceAI-splint* tool across: 15,244 *de novo* variants in genes linked to NDD in 7,221 participants; 839,150 variants from the complete set of *de novo* variants (see methods).

| **SpliceAI re-annotation Recommendation** | ***De novo* NDD variants (count / %)** | **All *de novo* variants in 100kGP (count / %)** |
| --- | --- | --- |
| **Total variants** | 15,244 | 838,921 |
| **Update** = large indel | 254 / 1.67% | 12,006 / 1.43% |
| **Update** = missing exon | 0 / 0% | 17 / <0.01% |
| **Update** = missing gene | 432 / 1.66% | 14,998 / 1.79% |
| **Update** = wrong REF | 0 / 0% | 38 / <0.01% |
| **Update** = possible splicing-effect at distance > 50 | 3,271 / 21.46% | 69,714 / 8.31% |
| **Update** = transcript boundary change | 516 / 3.38% | 18,143 / 2.16% |
| **TOTAL TO UPDATE** | 4,473 / 29.34% | 114,916 / 13.70% |
| **No recommended action** | 10,771 / 70.66% | 724,005 / 86.28% |

### Recommendations

In general, we recommend running all variants of interest using the SpliceAI command line tool with an up-to-date gene annotation file and increased distance. However, if working with a large set of variants without the computational resources to do so, we provide SpliceAI-splint to identify the subset of variants which would most benefit from reannotation. We recommend the following workflow for this use case:

1. Run Ensembl VEP with the SpliceAI plugin and annotate with precomputed scores as standard.
2. Run SpliceAI-splint over your variant set to identify the subset where the SpliceAI command line tool should be used in preference to the precomputed scores.
3. For variants SpliceAI-splint recommends reannotation for, run through the SpliceAI command line tool with an updated gene annotation file and increased distance.

This approach is intended to balance computational efficiency while maintaining accurate SpliceAI predictions.

## Discussion

The provision of precomputed SpliceAI scores has facilitated the identification of large numbers of candidate variants at a fraction of the computational time and cost of running spliceAI directly, however, these scores have several limitations which can reduce their accuracy in certain circumstances Here, we have highlighted ways in which splice altering variants may be both missed when using SpliceAI precomputed scores, and a schema whereby these ‘missed’ variants may subsequently be ‘rescued’. We report the expected additional yield of prioritised variants when using updated SpliceAI parameters and best bioinformatics practices to identify all predicted splice-altering variants using SpliceAI over large cohorts.

In this analysis we have identified the four main mechanisms by which candidate diagnostic variants may be missed when using precomputed scores: 1) updates to gene transcript annotations used by SpliceAI’s algorithm to inform prediction, 2) liftover errors introduced when converting precomputed scores between GRCh37 and GRCh38 3) large indels exceeding the size threshold for annotation using precomputed scores, and 4) increasing the distance within which potentially impacted splice sites are considered.

Besides genes completely missing from the annotation file, the largest source of outdated annotations came from changes to transcript boundaries, which likely results in the under annotation of variants that appear in close proximity to the termini of these transcripts. Although this may only represent a small number of variants per gene, it is important to highlight that these variants will most likely fall within the 5’ and 3’ UTRs, emerging contributors to human genetic disorders that have historically been under-explored(28). In addition to the findings presented here, we recently reported four pathogenic ClinVar variants in UTRs that were not captured using precomputed scores, but would be identified using spliceAI directly with updated transcript annotations(11). Amongst these was an additional variant whose predicted splice-altering impact couldn’t be identified using the precomputed scores due to a change in transcript boundaries (*SMARCB1*; chr22:23834262_C/T). In this case, the variant was within the boundaries of the transcript used in the original precomputed gene annotation file, and received a max Δ score of 0.08 in the precomputed scores. However when reassessed using the current MANE transcript annotation for the gene, which extends the transcript boundary by several thousand nucleotides, the max Δ score returned for the same variant is 0.97.

A key limitation of our study is that we have not been able to validate the splicing effect of the newly identified variants with max Δ >= 0.2, as the Genomics England participants with a newly identified candidate variant did not have RNA-seq data available. Whilst we acknowledge this represents a limitation in the ability to estimate the precision of our findings, SpliceAI predictions of splicing changes have been extensively validated by other studies(5,7,8,17). Of note, we found that the variants in the present study that were identified using an increased distance parameter had lower max Δ scores on average (Figure 1D) which may reflect that SpliceAI has less certainty when making predictions of variant effects at greater distance, however SpliceAI predictions at distances further than 50 nt have been validated in RNA studies elsewhere(7,17,19)

To reduce the burden of reanalysis, we have developed recommendations for the identification and reannotation of ‘missed’ variants, informed by our findings. We provide an open source tool ‘*spliceAI-splint*’ for use by the wider community to enable rapid reanalysis of variants, balancing computational efficiency with accuracy. Using this tool to predict candidate variants for reanalysis in the entire set of 15,244 *de novo* variants in 100kGP we identified 29% (4,473 variants) that may not have previously been correctly annotated using spliceAI precomputed scores alone, thus reducing the overall burden of a complete reannotation by ∼71%. Running spliceAI-splint on other genomic datasets we find the tool generally recommends reannotation of between 12 and 15% of variants.

Whilst a recomputed global set of spliceAI scores built on updated gene annotations and with increased distance would be of great use to the clinical genetics community, several caveats make it impractical. First, transcript and gene annotations are frequently updated, making it likely that any new set of precomputed scores would quickly become out of date again.

Additionally, it may only ever be feasible to precompute spliceAI predictions using a single representative transcript for each gene, as we have done in this study using MANE select transcripts. While the MANE select + clinical set is designed to encompass clinically relevant transcripts, it’s possible the analysis of predicted splicing changes on the multiple transcripts present for each gene could identify further splice-altering causes of diseases. This is compounded by limitations of current techniques for transcript annotation, specifically the reductive nature of 3’ UTR annotation(29,30). While ‘*spliceAI-splint*’ uses MANE v1.0 transcripts by default, users can adapt it to any set of transcripts, allowing for the expansion of SpliceAI re-annotation to encompass alternative non-canonical transcripts. We provide scripts and instructions to facilitate this in the Github repository.

A final consideration is that the provision of scores for all possible larger indels in human genes, would diminish the usability of the precomputed scores by expanding an already extensive set of data to the point of being prohibitively large. Taking into account all these considerations, and acknowledging that it is only a sizable minority of theoretical SNVs which are affected by annotation errors in the precomputed scores (8.34%, Figure 1C), we conclude that providing information on genomic regions and variant types where use of the precomputed scores is insufficient and instructions on how to supplement them is most appropriate at this time. We believe this approach balances the importance of accuracy in SpliceAI predictions with computational scalability and feasibility for clinical diagnostic teams.

Individuals affected by genetic conditions, particularly those that are rare, often face long diagnostic odysseys, with many failing to get a diagnosis within their lifetime. As genomic sequencing becomes more commonplace, and tools for the annotation of deleterious variants are improved or developed we are facing the need to return to individuals whose genomes have already been analysed in order to capture diagnoses that may previously have been missed(31). This reannotation places an enormous burden on clinical diagnostic teams who often have only limited resources to perform these reanalyses. It is therefore essential that any recommendations for reanalysis account for this burden, and mitigate the need for additional time, and compute resources where possible.

## Conclusions

Our study highlights the importance of using updated gene annotations and appropriate distance parameters when utilising SpliceAI, and the limitations of relying solely on precomputed scores. We provide best practices for employing precomputed scores and recommendations for cases where retrieving updated scores is necessary to help prioritise new candidate diagnostic variants. We identify areas where precomputed SpliceAI scores are most likely to need updating, and provide a tool for doing so, ensuring that these scores can continue to be effectively used in ever-growing cohorts. Our findings highlight the critical need for continuous updates and improvements in computational tools, and the provision of clear ‘best practice’ usage guidance, to keep pace with advancements in genomic research and annotation accuracy.

## Data Availability

All data from the Genomics England 100k Genomes Project was accessed via the Genomics England Trusted Research Environment (TRE) and is available to registered users of the National Genomics Research Library through the TRE platform. All other data are contained within the manuscript and associated repositories.

https://genome-euro.ucsc.edu/s/francois.leco/spliceai_precomputed_REF_errors_grch38

https://github.com/Computational-Rare-Disease-Genomics-WHG/spliceai-splint/tree/main

https://docs.google.com/spreadsheets/d/1Qt6hA7YGSmvvffXOyIZhopsPrptuLH7lbwcoVnrVDMA/edit?usp=sharing

## Data Availability

The code, and datasets supporting the conclusions of this article are available in the splice reannotation repository, DOI 10.5281/zenodo.16882891 and https://github.com/Computational-Rare-Disease-Genomics-WHG/spliceai-splint/tree/main. This repository includes data and scripts needed to produce spliceAI compatible transcript annotations, including; MANE Select V1.0 (GRCh38); And all scripts used to annotate variants that may be missed using precomputed SpliceAI scores. Additionally, we have made all data relating to REF mismatching in precomputed scores as a UCSC public session (https://genome-euro.ucsc.edu/s/francois.leco/spliceai_precomputed_REF_errors_grch38), with the full details for each gene included in supplementary table 1.

SpliceAI precomputed scores are available on the Illumina Basespace platform (https://basespace.illumina.com/s/5u6ThOblecrh ).

This research was made possible through access to data in the National Genomic Research Library, which is managed by Genomics England Limited (a wholly owned company of the Department of Health and Social Care). The National Genomic Research Library holds data provided by patients and collected by the NHS as part of their care and data collected as part of their participation in research. All data from the Genomics England 100k Genomes Project v15 (26.05.2022), including Genetic, phenotypic and RNA-seq data, was accessed via the Genomics England Trusted Research Environment (TRE) and is available to registered users of the National Genomics Research Library through the TRE platform.

## List of abbreviations

100kGP: Genomics England 100,000 Genomes Project
HPO: Human Phenotype Ontology
ICD10: International Statistical Classification of Diseases and Related Health Problems v10
MANE: Matched Annotation of NCBI and Ensembl
NDD: Neuro Developmental Disorder
SNV: Single Nucleotide Variant
UTR: Untranslated region
VEP: Variant effect predictor

## Declarations

## Acknowledgements

This research was made possible through access to data in the National Genomic Research Library, which is managed by Genomics England Limited (a wholly owned company of the Department of Health and Social Care). The National Genomic Research Library holds data provided by patients and collected by the NHS as part of their care and data collected as part of their participation in research. The National Genomic Research Library is funded by the National Institute for Health Research and NHS England. The Wellcome Trust, Cancer Research UK and the Medical Research Council have also funded research infrastructure.

We would like to acknowledge the Genomics England service desk, for the support and guidance they provided to make this work possible.

## Funding

RD is supported by grant funding from Novo Nordisk (to N.W.). NWhiffin and ACM-G are supported by a Sir Henry Dale Fellowship awarded to NWhiffin, jointly funded by the Wellcome Trust and the Royal Society (220134/Z/20/Z).

## Author Information

### Authors’ contributions

RD conceived and supervised the project, RD and ACM-G performed the analyses and co-wrote the manuscript. FL performed analyses and contributed to the manuscript. NW contributed to study design and provided feedback on the manuscript, SW contributed to clinical interpretation and provided feedback on the manuscript.

## Ethics Declarations

### Ethics approval and consent to participate

Ethics approval was granted by the HRA Committee East of England – Cambridge South (REC Ref 14/EE/1112). This research conforms to the principles of the Helsinki Declaration.All participants provided informed consent for their data to be part of the National Genomics Research Library and for use in research and publication.

### Consent for publication

All participants in this study have provided consent for their data to be part of the National Genomics Research Library and to the publication of research findings. For all cases, written informed consent for research use of clinical and genetic data was obtained from patients, their parents, or legal guardians in the case of those with intellectual disability.

### Competing interests

The authors declare no competing interests.

## Supplementary Data

Supplementary tables 1:5: Supplementary file 1

---

**Supplementary figure 1:**
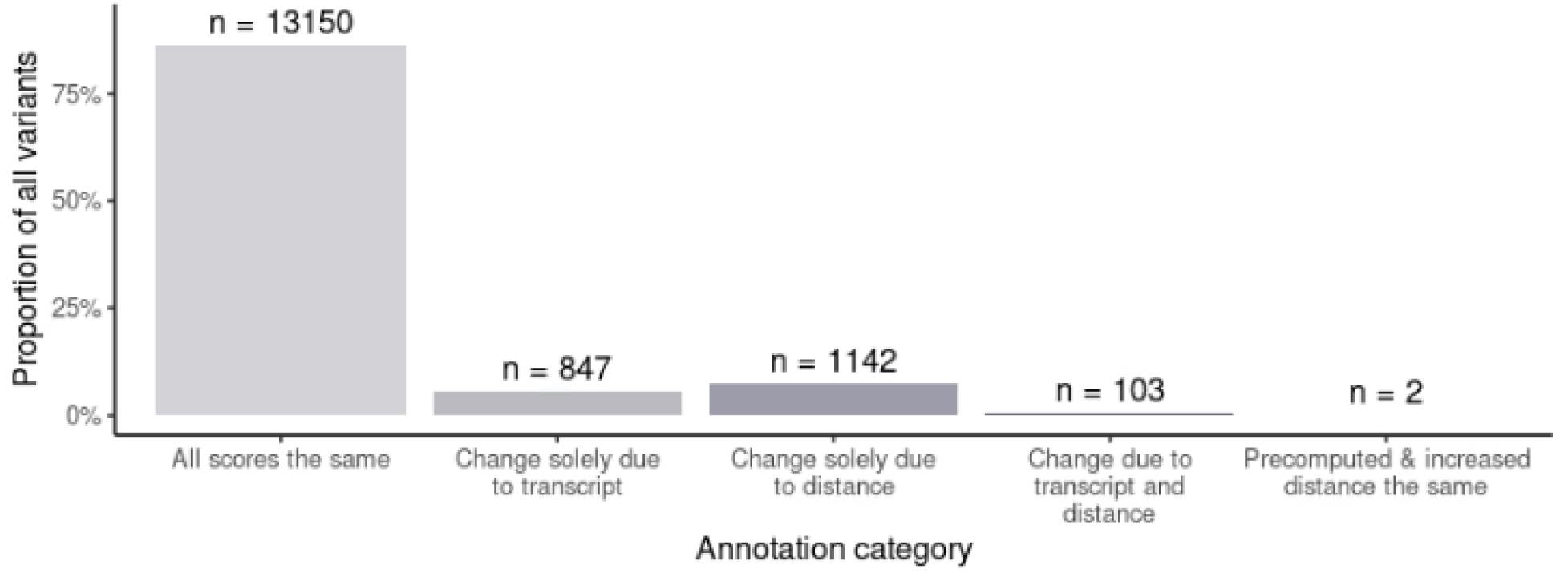
The landscape of change in SpliceAI scores between annotation runs. 86.26 percent of the full set of 15,244 *de novo* variants had no change in SpliceAI Max delta over each of the 3 conditions. 5.56% had a score change solely attributable to a change in transcript boundaries. 7.49% of variants’ scores were altered in response to a change in the distance parameter alone, and 0.68% of variants were impacted by *both* transcript annotation, *and* distance parameter changes. The scores of 2 variants initially changed in response to updated transcript annotations, but gained a new annotation that happened to be the same as their precomputed score scores when the distance parameter was increased.

**Supplementary figure 2:**
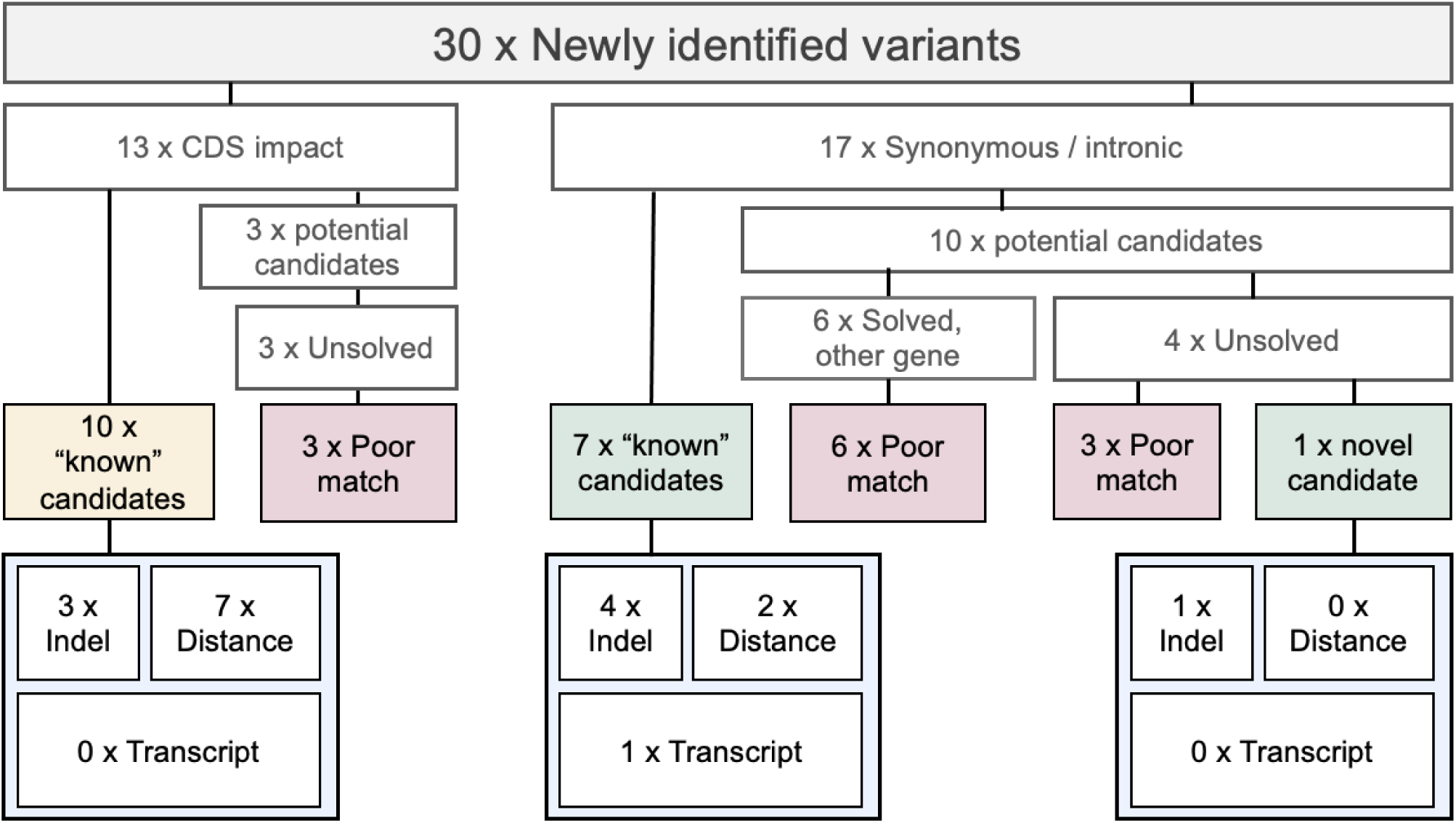
**Flowchart of variant interpretation for the 30 newly identified variants from 100kGP**, including the mechanism by which they were detected. Variants that were not considered good candidates for the individual’s phenotype, or that were also predicted to have a ‘coding effect’ are shown in pink and yellow respectively, candidate diagnostic variants with a likely splice altering effect are shown in green. The mechanisms by which each of the potentially diagnostic candidate variants were detected are shown in pale blue.

## Notes

### Competing Interest Statement

The authors have declared no competing interest.

### Author Declarations

Ethics committee/IRB of the HRA Committee East of England, Cambridge South (REC Ref 14/EE/1112) gave ethical approval for this work.

